# Blood-Based Transcriptomic and Proteomic Biomarkers of Emphysema

**DOI:** 10.1101/2022.10.25.22281458

**Authors:** Rahul Suryadevara, Andrew Gregory, Robin Lu, Zhonghui Xu, Aria Masoomi, Sharon M. Lutz, Seth Berman, Jeong H. Yun, Aabida Saferali, Craig P. Hersh, Edwin K. Silverman, Jennifer Dy, Katherine A. Pratte, Russell P. Bowler, Peter J. Castaldi, Adel Boueiz, the COPDGene investigators

**Author notes:** **Corresponding Author:** Adel Boueiz, Channing Division of Network Medicine, Brigham and Women’s Hospital, 181 Longwood Avenue, Boston, MA, 02115. Equal contribution (Peter J. Castaldi, Adel Boueiz). Authors’ email addresses: Rahul Suryadevara, Andrew Gregory, Robin Lu, Zhonghui Xu, Aria Masoomi, Sharon M. Lutz, Seth Berman, Jeong H. Yun, Aabida Saferali, Craig P. Hersh, Edwin K. Silverman, Jennifer Dy, Katherine A. Pratte, Russell P. Bowler, Peter J. Castaldi, Adel Boueiz. **Author Contributions:** Drs. Boueiz and Castaldi had full access to all the data in the study, take responsibility for the integrity of the data and the accuracy of the data analysis, had authority over manuscript preparation and the decision to submit the manuscript for publication. *Study concept and design:* Boueiz, Castaldi, Xu *Acquisition, analysis, or interpretation of data:* All authors *Drafting of the manuscript:* Suryadevara, Boueiz, Castaldi *Critical revision of the manuscript for important intellectual content:* All authors *Statistical analysis:* Xu, Lutz, Castaldi, Boueiz *Obtained funding:* Boueiz, Castaldi, Silverman *Study supervision:* All authors All authors gave final approval of the version to be published. **This article has an online data supplement, which is accessible from this issue’s table of content online at www.atjsjournals.org**.

## Abstract

**Rationale:** Emphysema is a COPD phenotype with important prognostic implications. Identifying blood-based biomarkers of emphysema will facilitate early diagnosis and development of targeted therapies.

**Objectives:** Discover blood omics biomarkers for chest CT-quantified emphysema and develop predictive biomarker panels.

**Methods:** Emphysema blood biomarker discovery was performed using differential gene expression, alternative splicing, and protein association analyses in a training set of 2,370 COPDGene participants with available whole blood RNA sequencing, plasma SomaScan proteomics, and clinical data. Validation was conducted in a testing set of 1,016 COPDGene subjects. Since low body mass index (BMI) and emphysema often co-occur, we performed a mediation analysis to quantify the effect of BMI on gene and protein associations with emphysema. Elastic net models were also developed in the training sample sequentially using clinical, complete blood count (CBC) cell proportions, RNA sequencing, and proteomic biomarkers to predict quantitative emphysema. Model accuracy was assessed in the testing sample by the area under the receiver-operator-characteristic-curves (AUROC) for subjects stratified into tertiles of emphysema severity.

**Measurements and Main Results:** 4,913 genes, 1,478 isoforms, 386 exons, and 881 proteins were significantly associated with emphysema *(FDR 10%)* and yielded 109 biological pathways. 75% of the genes and 77% of the proteins associated with emphysema showed evidence of mediation by BMI. The highest-performing predictive model used clinical, CBC, and protein biomarkers, distinguishing the top from the bottom tertile of emphysema with an AUROC of 0.92.

**Conclusions:** Blood transcriptome and proteome-wide analyses reveal key biological pathways of emphysema and enhance the prediction of emphysema.

**AT A GLANCE COMMENTARY:** *Scientific Knowledge on the Subject:* Differential gene expression and protein analyses have uncovered some of the molecular underpinnings of emphysema. However, no studies have assessed alternative splicing mechanisms and analyzed proteomic data from recently developed high-throughput panels. In addition, although emphysema has been associated with low body mass index (BMI), it is still unclear how BMI affects the transcriptome and proteome of the disease. Finally, the effectiveness of multi-omic biomarkers in determining the severity of emphysema has not yet been investigated.

*What This Study Adds to the Field:* We performed whole-blood genome-wide RNA sequencing and plasma SomaScan proteomic analyses in the large and well-phenotyped COPDGene study. In addition to confirming earlier findings, our differential gene expression, alternative splicing, and protein analyses identified novel biomarkers and pathways of chest CT-quantified emphysema. Our mediation analysis detected varying degrees of transcriptomic and proteomic mediation due to BMI. Our supervised machine learning modeling demonstrated the utility of incorporating multi-omics data in enhancing the prediction of emphysema.

## INTRODUCTION

Chronic obstructive pulmonary disease (COPD) is a leading cause of morbidity and mortality (1). Emphysema, the anatomic destruction of lung parenchyma frequently observed in COPD subjects, has been independently associated with an increased risk for cardiovascular disease, lung cancer, and mortality (2-4). Timely diagnosis calls for a blood-based predictive model as it may identify emphysema in subjects where computed tomography (CT) scans are not clinically indicated. Emphysema blood biomarkers would also overcome the issues of radiation exposure and false positive findings associated with CT scans (5). In addition, early disease biomarkers and a stronger understanding of the molecular basis of emphysema are needed to develop novel personalized therapies to improve the prognosis of affected individuals (2, 6, 7).

Previous transcriptomic studies have identified emphysema-associated genes (such as *COL6A1, CD19, PTX3*, and *RAGE*) and biological processes (such as innate and adaptive immunity, inflammation, and tissue remodeling) primarily from gene expression analyses using lung tissue samples (8-12). However, fewer studies have evaluated the associations of emphysema with blood transcriptomics, alternative splicing, or proteomics. Alternative splicing, the regulatory process in which multi-exon human genes are expressed in multiple transcript isoforms, has been implicated in the pathophysiology of several lung diseases such as asthma, pulmonary fibrosis, pulmonary arterial hypertension, and COPD (13-19). Protein levels have also been studied for potential emphysema biomarker identification, and sRAGE, ICAM1, CCL20, and adiponectin levels in blood and eotaxin levels in bronchoalveolar lavage fluid were found to be associated with emphysema (20-23), though the protein panels used for these studies included fewer proteins than the more recently developed panels. Finally, previous research that used blood-based emphysema predictive models had small sample sizes and only tested one ‘omic modality at a time (20, 24-27).

We hypothesized that 1) transcriptomic and proteomic characterization of smokers would elucidate emphysema pathobiology and yield novel disease biomarkers, 2) many emphysema associations with transcripts and proteins are influenced by BMI, and 3) multi-omic modeling would provide improved prediction of emphysema relative to readily available clinical variables. To test these hypotheses, we analyzed whole-blood genome-wide RNA sequencing (RNA-seq) and plasma SomaScan proteomic data from the large and well-phenotyped COPDGene study. Given the high clinical correlation between emphysema and BMI (28), we performed mediation analysis to understand the influence of BMI on emphysema-associated genes and proteins. We also developed machine learning predictive models for emphysema using transcriptomic and proteomic biomarkers. Some of these results have been previously reported as an abstract (29) and a preprint (30).

## METHODS

### Study description

Participants were recruited from the COPDGene study (NCT00608764, www.copdgene.org), a longitudinal study investigating the genetic basis of COPD. The COPDGene population consists of 10,371 non-Hispanic white and African American subjects, 44-90 years old with an average of 44 pack-years of lifetime cigarette smoking history (31). Subjects had varying degrees of COPD severity, as measured by the Global Initiative for Chronic Obstructive Lung Disease (GOLD) grading system. COPDGene obtained 5-year follow-up data and is currently obtaining 10-year follow-up data of available subjects. Questionnaires, chest CT scans, and spirometry have been gathered at 21 clinical facilities in the United States. RNA-seq and plasma proteomic measurements were obtained from a subset of subjects at their 5-year follow-up visit (Visit 2). Each center acquired institutional review board approval and written informed consents. In our analyses, we used the COPDGene Visit 2 data, which included 3,386 subjects with available clinical, RNA-seq, and SomaScan proteomic data.

### Emphysema quantification

Using the Thirona software (www.thirona.eu), emphysema was quantified as the 15^th^ percentile of the attenuation histogram + 1,000 Hounsfield units (HU), corrected for the inspiratory depth variations using Multi-Ethnic Study of Atherosclerosis normative equations (predicted lung volume using baseline age, time-varying height, and BMI) (adjusted Perc15 density) (32-34). This correction was made since it had been demonstrated to provide a more robust measure of longitudinal changes in emphysema (33).

### Training and testing samples

We randomly partitioned our studied cohort into training and testing samples comprising 70% and 30% of the subjects, respectively. All association and mediation analyses, as well as prediction model training, were conducted using the training sample. The validation of the identified biomarkers and constructed predictive model was carried out in the testing sample.

### RNA isolation, library preparation, filtering, and normalization

Illumina sequencers were utilized to obtain gene, isoform, and exon counts from total blood RNA isolated from Visit 2 participants. Genomic features with very low expression (average counts per million (CPM) < 0.2 or number of subjects with CPM < 0.5 less than 50) or extremely highly expressed genes (number of subjects with CPM > 50,000 less than 50) were filtered out prior to applying trimmed mean of M values normalization from edgeR (v3.24.3), which accounts for differences in sequencing depth (35). Counts were transformed to log2 CPM values and quantile-normalized to further remove systematic noise from the data.

### Protein measurements and filtering

At Visit 2, plasma samples were assayed for 4,979 proteins using the SomaScan Human Plasma 5.0K assay, a multiplex aptamer-based assay (SomaLogic, Boulder, Colorado) (36). The SomaScan data was standardized per the SomaLogic protocol to control for inter-assay variation between analytes and batch differences between plates (37). Samples with low volume, failed hybridization control, or failed dilution scale were removed. Proteomic data for 5,670 participants passed quality control. The protein counts were transformed to log2 RFU (relative fluorescent units) values.

### RNA-seq differential expression, usage, and protein association analyses

We used the limma-voom linear modeling approach (as implemented in limma v3.38.3) to test for the associations between emphysema and whole blood RNA transcripts (38, 39). The diffSplice function from limma was used to test for differential usage of isoforms and exons. While differential expression refers to the change in the *absolute* expression levels of a feature, differential usage captures alternative splicing and refers to the change in the *relative* expression levels of the isoforms/exons within a given gene. The associations of the SomaScan proteins with emphysema were tested using multivariable linear modeling. In the emphysema “primary” model, we adjusted for age, race, sex, pack-years of smoking, current smoking status, forced expiratory volume in one second (FEV_1_), complete blood count (CBC) cell proportions, CT scanner model, and library preparation batch for RNA-seq or clinical center for proteins. The validation rate in the testing sample was determined based on a threshold P-value < 0.1 and a consistent direction of effect in the training and testing datasets. A sensitivity analysis was performed in which the list of covariates from the primary model was expanded to include BMI. To select biomarkers for inclusion in the prediction model, we ran additional models only adjusted for the technical factors (CT scanner model and library preparation batch for RNA-seq or clinical center for proteins). Multiple comparisons were corrected with the Benjamini-Hochberg method using a threshold of significance of a false discovery rate (FDR) of 10% (40).

### Mediation analysis

We conducted mediation analysis using the medflex R package (41) to distinguish how much of the effect of emphysema on gene expression or protein levels acted through BMI (referred to as the indirect effect) and how much of the effect of emphysema directly influenced gene expression or protein levels (referred to as the direct effect). The analysis was performed on the significant genes and proteins identified in the primary association analysis. A mediated proportion representing the ratio of the indirect effect over the total effect was computed for each gene with significant total effect. The P-values of the direct, indirect, and total effects for each biomarker were subject to a threshold significance of 10% FDR.

### Gene set enrichment analyses

The biological enrichment of the gene sets derived from the gene expression, transcript usage, and protein association analyses was evaluated using the topGO (v2.33.1) weight01 algorithm, which accounts for the dependency in the Gene Ontology (GO) topology (42). We only reported GO pathways with at least three significant genes and an adjusted P-value < 0.005.

### Predictive modeling

We constructed elastic net models to predict cross-sectional emphysema (43). The outcome variable was the adjusted Perc15 density. We utilized clinical variables that are readily available in the primary care setting (age, race, sex, BMI, pack-years of smoking, and current smoking status), CBC (proportions of neutrophils, eosinophils, monocytes, lymphocytes, and platelets), and the RNA-seq and proteins that reached statistical significance in the association analyses performed in the training data (adjusted only for the scanner model and library preparation batch or clinical center). To determine the top performing RNA data type to be used in the main models, we first constructed models using clinical + gene, clinical + isoform, and clinical + exon counts. We then constructed models in the following order: clinical only, clinical + CBC, clinical + CBC + RNA-seq, clinical + CBC + proteins, and clinical + CBC + RNA-seq + proteins. The outcome and the predictors were centered and scaled. The models were trained using 10-fold cross-validation, minimizing the mean squared error (44) on the left-out fold. After model training on the continuous emphysema variable, we classified subjects into tertiles of adjusted Perc15 density. We evaluated the predictive performances of the models in the testing sample using R^2^ for the continuous emphysema and the area-under-receiver-operator-characteristic curve (AUROC) for the model accuracy to distinguish subjects in the highest and lowest tertiles of emphysema severity. We compared the AUROCs with the DeLong test using the pROC R package (45). Finally, predictors were ranked by the absolute values of their coefficients from the regression model.

### Statistical analysis

Data were reported as means with standard deviations or counts with percentages. Continuous variables were tested with Kruskal-Wallis and categorical variables were tested with chi-square. Upregulated versus downregulated genes as well as positive versus negative signs of the protein coefficients are provided with respect to their relationships with adjusted Perc15 density (i.e., negative coefficients indicate a greater extent of emphysema).

Additional methods are available in the Supplement.

## RESULTS

### Subject characteristics

3,386 subjects from COPDGene Visit 2 with complete clinical, RNA-seq, and protein data were included in our analyses (Figure 1). As shown in Table 1, the included subjects were mostly non-Hispanic whites with a balanced representation by sex, a mean age of 65, a mean BMI of 29, and a mean of 41 pack-years of smoking. The subjects’ characteristics did not significantly differ between the training and testing data, which consisted of 2,370 and 1,016 subjects, respectively. A comparison of subjects with and without missing data showed that the two groups were largely similar (Table E1). A schematic overview of the analyses performed is illustrated in Figure E1.

**Figure 1.**
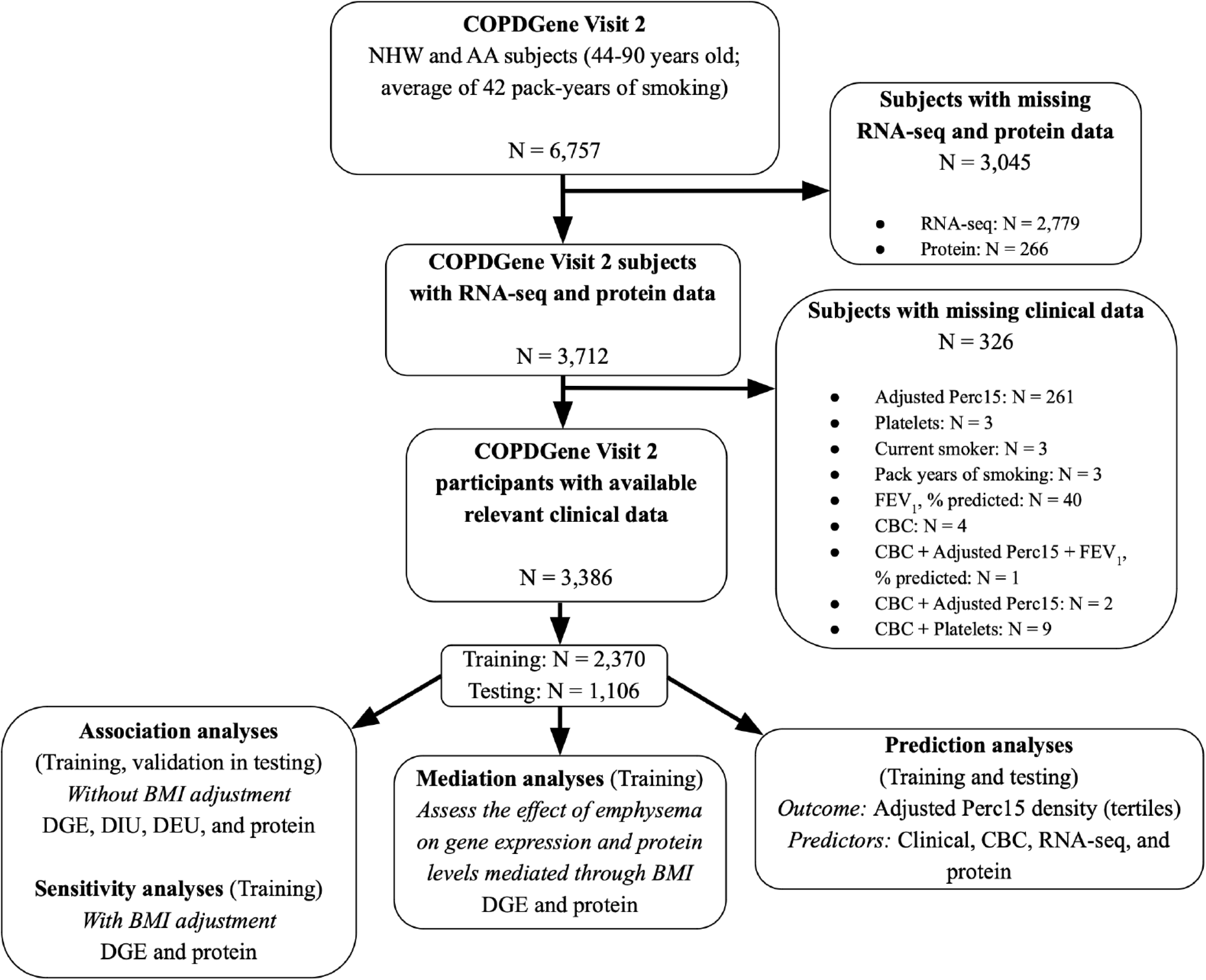
COPDGene Visit 2 participant flow diagram. Abbreviations: AA = African American, CBC = Complete blood count, DGE = Differential gene expression, DIU = Differential isoform usage, DEU = Differential exon usage, FEV_1_ = Forced expiratory volume, NHW = Non-Hispanic White.

**Table 1.**
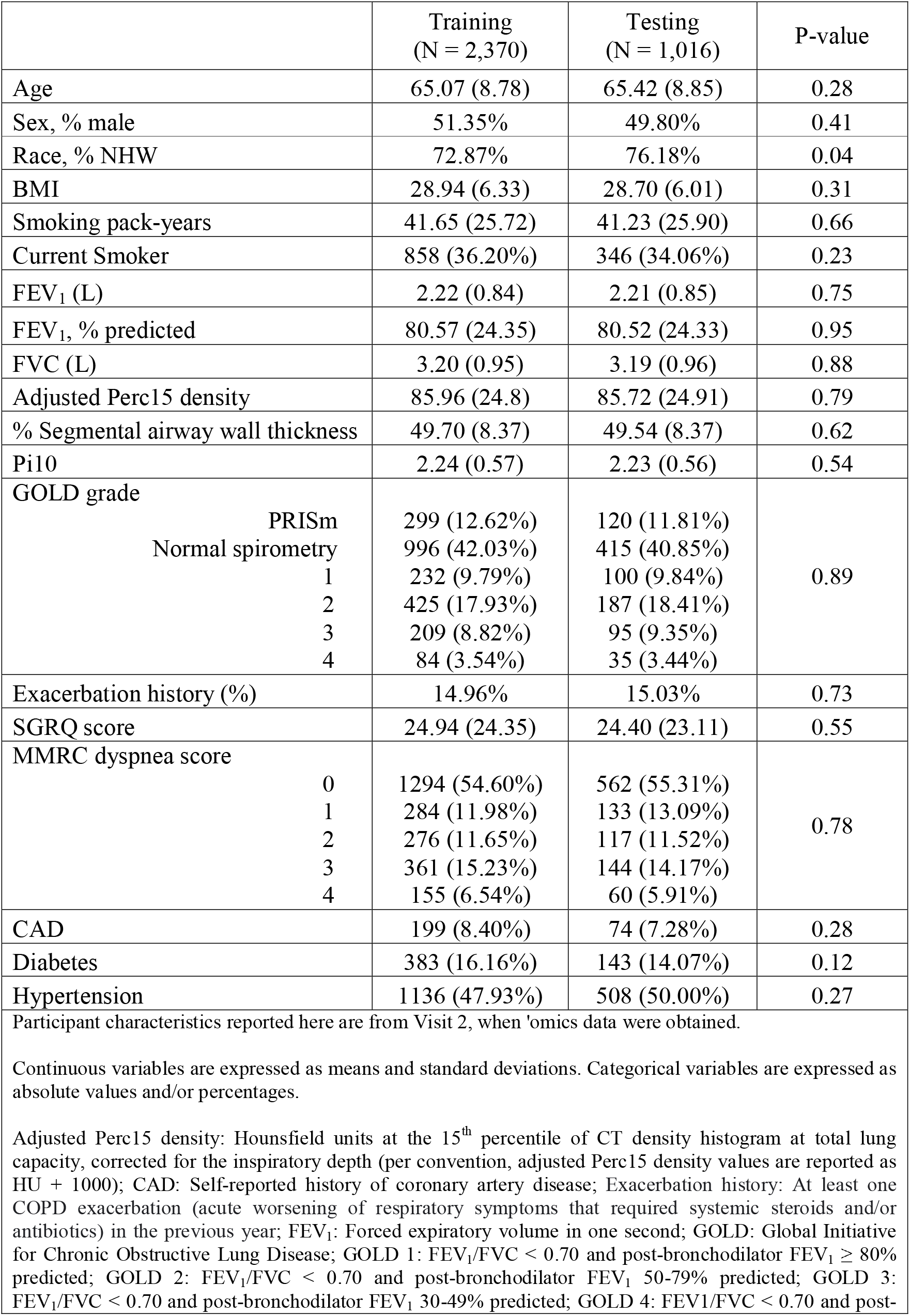
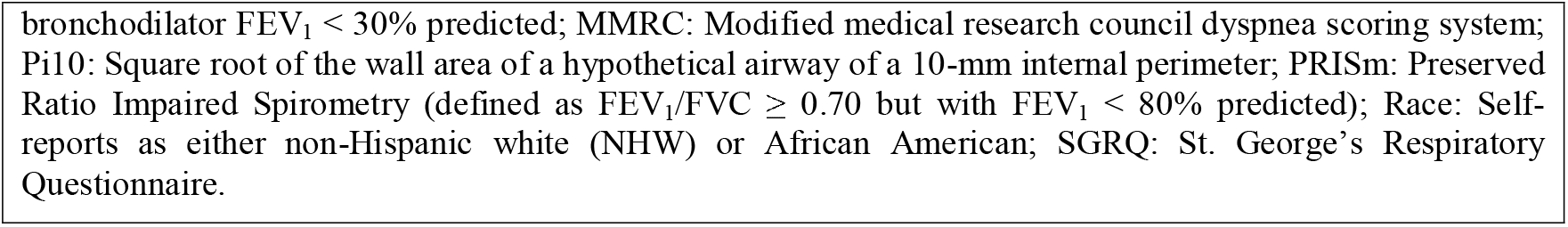
Characteristics of subjects in the training and testing datasets in COPDGene Visit 2.

|  | Training<br>(N = 2,370) | Testing<br>(N = 1,016) | P-value |
| --- | --- | --- | --- |
| Age | 65.07 (8.78) | 65.42 (8.85) | 0.28 |
| Sex, % male | 51.35% | 49.80% | 0.41 |
| Race, % NHW | 72.87% | 76.18% | 0.04 |
| BMI | 28.94 (6.33) | 28.70 (6.01) | 0.31 |
| Smoking pack-years | 41.65 (25.72) | 41.23 (25.90) | 0.66 |
| Current Smoker | 858 (36.20%) | 346 (34.06%) | 0.23 |
| FEV<sub>1</sub> (L) | 2.22 (0.84) | 2.21 (0.85) | 0.75 |
| FEV<sub>1</sub>, % predicted | 80.57 (24.35) | 80.52 (24.33) | 0.95 |
| FVC (L) | 3.20 (0.95) | 3.19 (0.96) | 0.88 |
| Adjusted Perc15 density | 85.96 (24.8) | 85.72 (24.91) | 0.79 |
| % Segmental airway wall thickness | 49.70 (8.37) | 49.54 (8.37) | 0.62 |
| Pi10 | 2.24 (0.57) | 2.23 (0.56) | 0.54 |
| GOLD grade |  |  |  |
| PRISm | 299 (12.62%) | 120 (11.81%) |  |
| Normal spirometry | 996 (42.03%) | 415 (40.85%) |  |
| 1 | 232 (9.79%) | 100 (9.84%) | 0.89 |
| 2 | 425 (17.93%) | 187 (18.41%) |  |
| 3 | 209 (8.82%) | 95 (9.35%) |  |
| 4 | 84 (3.54%) | 35 (3.44%) |  |
| Exacerbation history (%) | 14.96% | 15.03% | 0.73 |
| SGRQ score | 24.94 (24.35) | 24.40 (23.11) | 0.55 |
| MMRC dyspnea score |  |  |  |
| 0 | 1294 (54.60%) | 562 (55.31%) |  |
| 1 | 284 (11.98%) | 133 (13.09%) | 0.78 |
| 2 | 276 (11.65%) | 117 (11.52%) |  |
| 3 | 361 (15.23%) | 144 (14.17%) |  |
| 4 | 155 (6.54%) | 60 (5.91%) |  |
| CAD | 199 (8.40%) | 74 (7.28%) | 0.28 |
| Diabetes | 383 (16.16%) | 143 (14.07%) | 0.12 |
| Hypertension | 1136 (47.93%) | 508 (50.00%) | 0.27 |
| Participant characteristics reported here are from Visit 2, when 'omics data were obtained. |  |  |  |
| Continuous variables are expressed as means and standard deviations. Categorical variables are expressed as absolute values and/or percentages. |  |  |  |
| Adjusted Perc15 density: Hounsfield units at the 15<sup>th</sup> percentile of CT density histogram at total lung capacity, corrected for the inspiratory depth (per convention, adjusted Perc15 density values are reported as HU + 1000); CAD: Self-reported history of coronary artery disease; Exacerbation history: At least one COPD exacerbation (acute worsening of respiratory symptoms that required systemic steroids and/or antibiotics) in the previous year; FEV<sub>1</sub>: Forced expiratory volume in one second; GOLD: Global Initiative for Chronic Obstructive Lung Disease; GOLD 1: FEV<sub>1</sub>/FVC < 0.70 and post-bronchodilator FEV<sub>1</sub> ≥ 80% predicted; GOLD 2: FEV<sub>1</sub>/FVC < 0.70 and post-bronchodilator FEV<sub>1</sub> 50-79% predicted; GOLD 3: FEV<sub>1</sub>/FVC < 0.70 and post-bronchodilator FEV<sub>1</sub> 30-49% predicted; GOLD 4: FEV<sub>1</sub>/FVC < 0.70 and post- |  |  |  |

bronchodilator $FEV_1 < 30\%$ predicted; MMRC: Modified medical research council dyspnea scoring system; Pi10: Square root of the wall area of a hypothetical airway of a 10-mm internal perimeter; PRISm: Preserved Ratio Impaired Spirometry (defined as $FEV_1/FVC \geq 0.70$ but with $FEV_1 < 80\%$ predicted); Race: Self-reports as either non-Hispanic white (NHW) or African American; SGRQ: St. George's Respiratory Questionnaire.

### Differential gene expression analysis

We performed differential gene expression (DGE) analysis in the 2,370 subjects of the training dataset. 4,913 out of 19,177 genes reached significance at 10% FDR (Table 2 and E2). 2,339 genes were up-regulated and 2,574 were down-regulated with respect to adjusted Perc15 density (i.e., they have opposite directions for their associations with emphysema) (Figure 2A). The GO enrichment analysis identified 44 significantly enriched biological processes, including neutrophil degranulation, regulation of NF-κB signaling, viral transcription, T cell proliferation, and regulation of TNF-mediated signaling (Tables 3 and E3).

**Table 2.**
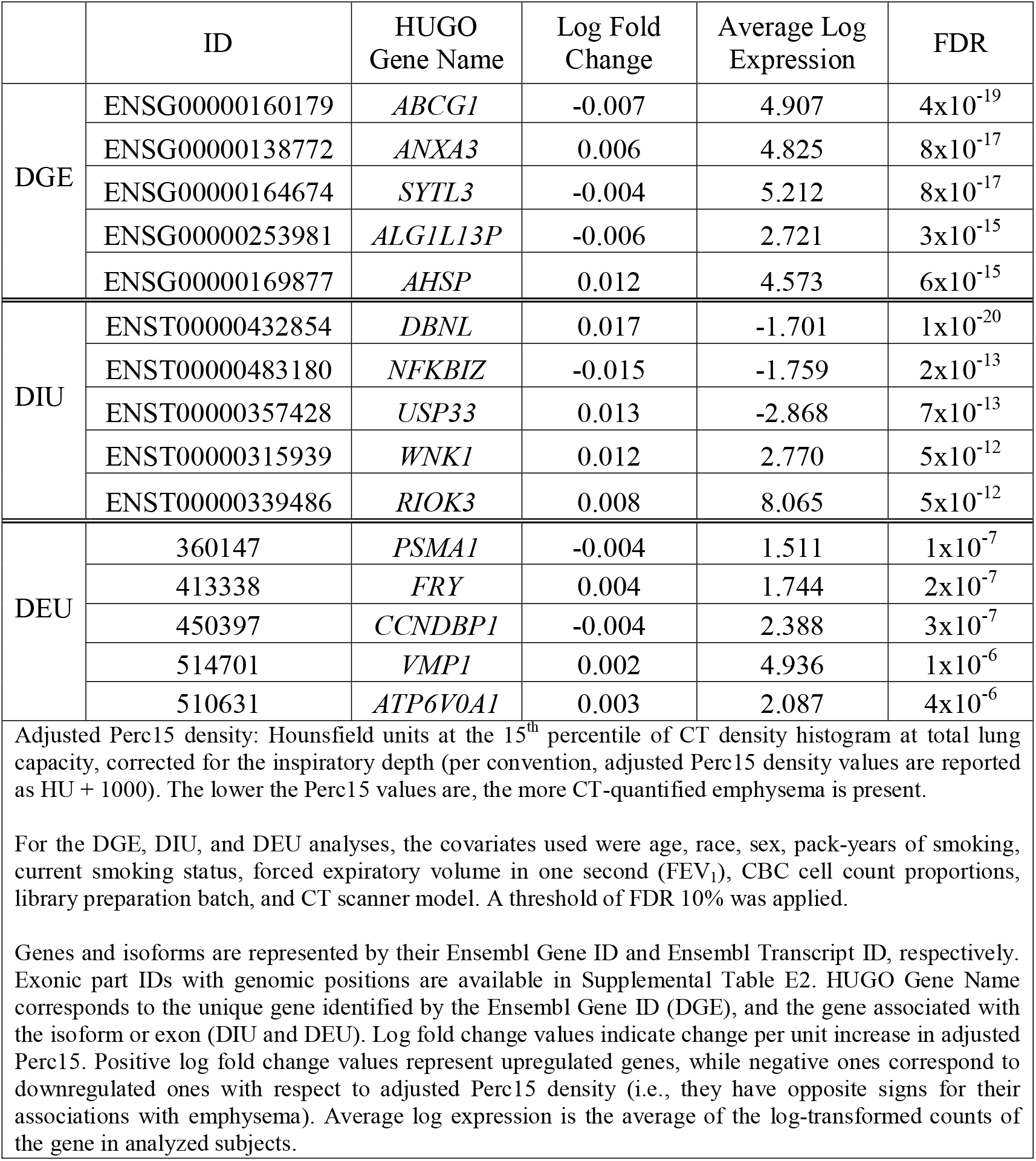
Top 5 differentially expressed genes (DGE), differentially used isoforms (DIU), and differentially used exons (DEU) associated to adjusted Perc15 density.

|  | ID | HUGO Gene Name | Log Fold Change | Average Log Expression | FDR |
| --- | --- | --- | --- | --- | --- |
| DGE | ENSG00000160179 | *ABCG1* | -0.007 | 4.907 | $4 \times 10^{-19}$ |
| | ENSG00000138772 | *ANXA3* | 0.006 | 4.825 | $8 \times 10^{-17}$ |
| | ENSG00000164674 | *SYTL3* | -0.004 | 5.212 | $8 \times 10^{-17}$ |
| | ENSG00000253981 | *ALG1L13P* | -0.006 | 2.721 | $3 \times 10^{-15}$ |
| | ENSG00000169877 | *AHSP* | 0.012 | 4.573 | $6 \times 10^{-15}$ |
| DIU | ENST00000432854 | *DBNL* | 0.017 | -1.701 | $1 \times 10^{-20}$ |
| | ENST00000483180 | *NFKBIZ* | -0.015 | -1.759 | $2 \times 10^{-13}$ |
| | ENST00000357428 | *USP33* | 0.013 | -2.868 | $7 \times 10^{-13}$ |
| | ENST00000315939 | *WNK1* | 0.012 | 2.770 | $5 \times 10^{-12}$ |
| | ENST00000339486 | *RIOK3* | 0.008 | 8.065 | $5 \times 10^{-12}$ |
| DEU | 360147 | *PSMA1* | -0.004 | 1.511 | $1 \times 10^{-7}$ |
| | 413338 | *FRY* | 0.004 | 1.744 | $2 \times 10^{-7}$ |
| | 450397 | *CCNDBP1* | -0.004 | 2.388 | $3 \times 10^{-7}$ |
| | 514701 | *VMP1* | 0.002 | 4.936 | $1 \times 10^{-6}$ |
| | 510631 | *ATP6V0A1* | 0.003 | 2.087 | $4 \times 10^{-6}$ |
| Adjusted Perc15 density: Hounsfield units at the 15<sup>th</sup> percentile of CT density histogram at total lung capacity, corrected for the inspiratory depth (per convention, adjusted Perc15 density values are reported as HU + 1000). The lower the Perc15 values are, the more CT-quantified emphysema is present. |  |  |  |  |  |
| For the DGE, DIU, and DEU analyses, the covariates used were age, race, sex, pack-years of smoking, current smoking status, forced expiratory volume in one second (FEV<sub>1</sub>), CBC cell count proportions, library preparation batch, and CT scanner model. A threshold of FDR 10% was applied. |  |  |  |  |  |
| Genes and isoforms are represented by their Ensembl Gene ID and Ensembl Transcript ID, respectively. Exonic part IDs with genomic positions are available in Supplemental Table E2. HUGO Gene Name corresponds to the unique gene identified by the Ensembl Gene ID (DGE), and the gene associated with the isoform or exon (DIU and DEU). Log fold change values indicate change per unit increase in adjusted Perc15. Positive log fold change values represent upregulated genes, while negative ones correspond to downregulated ones with respect to adjusted Perc15 density (i.e., they have opposite signs for their associations with emphysema). Average log expression is the average of the log-transformed counts of the gene in analyzed subjects. |  |  |  |  |  |

**Table 3.** Selected top 10 gene ontology (GO) biological processes enriched in differentially expressed genes (DGE), differentially used isoforms (DIU), and differentially used exons (DEU) associated to adjusted Perc15 density. GO terms were selected based on potential biological relevance to emphysema.

|  | GO.ID | GO Term | Total number of genes in category | Number of adjusted Perc15 density-associated genes in category | Adjusted P-value |
| --- | --- | --- | --- | --- | --- |
| DGE | GO:0006614 | SRP-dependent cotranslational protein targeting to membrane | 99 | 72 | $2 \times 10^{-21}$ |
| | GO:0006413 | Translational initiation | 185 | 97 | $2 \times 10^{-19}$ |
| | GO:0000184 | Nuclear-transcribed mRNA catabolic process, nonsense-mediated decay | 120 | 77 | $5 \times 10^{-14}$ |
| | GO:0019083 | Viral transcription | 174 | 87 | $3 \times 10^{-13}$ |
| | GO:0043312 | Neutrophil degranulation | 466 | 212 | $9 \times 10^{-12}$ |
| | GO:0002181 | Cytoplasmic translation | 98 | 44 | $7 \times 10^{-7}$ |
| | GO:0051092 | Positive regulation of NF-kappaB transcription factor activity | 144 | 70 | $3 \times 10^{-6}$ |
| | GO:0046718 | Viral entry into host cell | 111 | 58 | $6 \times 10^{-6}$ |
| | GO:0042102 | Positive regulation of T cell proliferation | 84 | 47 | $1 \times 10^{-5}$ |
| | GO:0010803 | Regulation of tumor necrosis factor-mediated signaling pathway | 51 | 25 | $2 \times 10^{-5}$ |
| DIU | GO:0006413 | Translational initiation | 176 | 35 | $2 \times 10^{-5}$ |
| | GO:0045070 | Positive regulation of viral genome replication | 32 | 13 | $3 \times 10^{-5}$ |
| | GO:0043044 | ATP-dependent chromatin remodeling | 63 | 13 | $7 \times 10^{-5}$ |
| | GO:0090263 | Positive regulation of canonical WNT signaling pathway | 107 | 26 | $7 \times 10^{-5}$ |
| | GO:0018105 | Peptidyl-serine phosphorylation | 209 | 45 | $1 \times 10^{-4}$ |
| | GO:0032092 | Positive regulation of protein binding | 59 | 17 | $1 \times 10^{-4}$ |
| | GO:0006614 | SRP-dependent cotranslational protein targeting to membrane | 94 | 23 | $2 \times 10^{-4}$ |
| | GO:0000184 | Nuclear-transcribed mRNA catabolic process, nonsense-mediated decay | 118 | 28 | $2 \times 10^{-4}$ |
| | GO:0090263 | Positive regulation of transcription by RNA polymerase II | 690 | 106 | $3 \times 10^{-4}$ |
| | GO:0019083 | Viral transcription | 172 | 31 | $5 \times 10^{-4}$ |
| | GO:0019079 | Viral genome replication | 103 | 23 | $5 \times 10^{-4}$ |
| DEU | GO:0006413 | Translational initiation | 181 | 13 | $1 \times 10^{-4}$ |
|  | GO:0006995 | Cellular response to nitrogen starvation | 9 | 3 | 0.001 |
|  | GO:1904667 | Negative regulation of ubiquitin protein ligase activity | 9 | 3 | 0.001 |
|  | GO:1901991 | Negative regulation of mitotic cell cycle phase transition | 182 | 8 | 0.002 |
|  | GO:0006614 | SRP-dependent cotranslational protein targeting to membrane | 94 | 8 | 0.002 |
|  | GO:0000422 | Autophagy of mitochondrion | 72 | 8 | 0.002 |
|  | GO:0071560 | Cellular response to transforming growth factor beta stimulus | 133 | 9 | 0.002 |
|  | GO:0045722 | Positive regulation of gluconeogenesis | 11 | 3 | 0.002 |
|  | GO:0043124 | Negative regulation of I-kappaB kinase/NF-kappaB signaling | 38 | 5 | 0.002 |
|  | GO:0050821 | Protein stabilization | 142 | 10 | 0.002 |
| Adjusted Perc15 density: Hounsfield units at the 15<sup>th</sup> percentile of CT density histogram at total lung capacity, corrected for the inspiratory depth (per convention, adjusted Perc15 density values are reported as HU + 1000). The lower the Perc15 values are, i.e., the closer to -1,000 HU, the more CT-quantified emphysema is present.<br>For the DGE, DIU, and DEU analyses, covariates used were age, race, sex, pack-years of smoking, current smoking status, forced expiratory volume in one second (FEV<sub>1</sub>), CBC cell count proportions, library preparation batch, and CT scanner model.<br>We only reported the GO pathways with at least 3 significant genes. Enriched GO terms were identified using the weighted Fisher's test P-values < 0.005. Selected GO terms with the lowest P-values in the DGE, DIU, and DEU analyses are listed. Total number of genes in category refers to all genes studied that fall under the GO term. The number of adjusted Perc15-associated genes in category refers to the genes that reached significance (*FDR 10%*) in the DGE, DIU, and DEU analyses. |  |  |  |  |  |

**Table 4.**
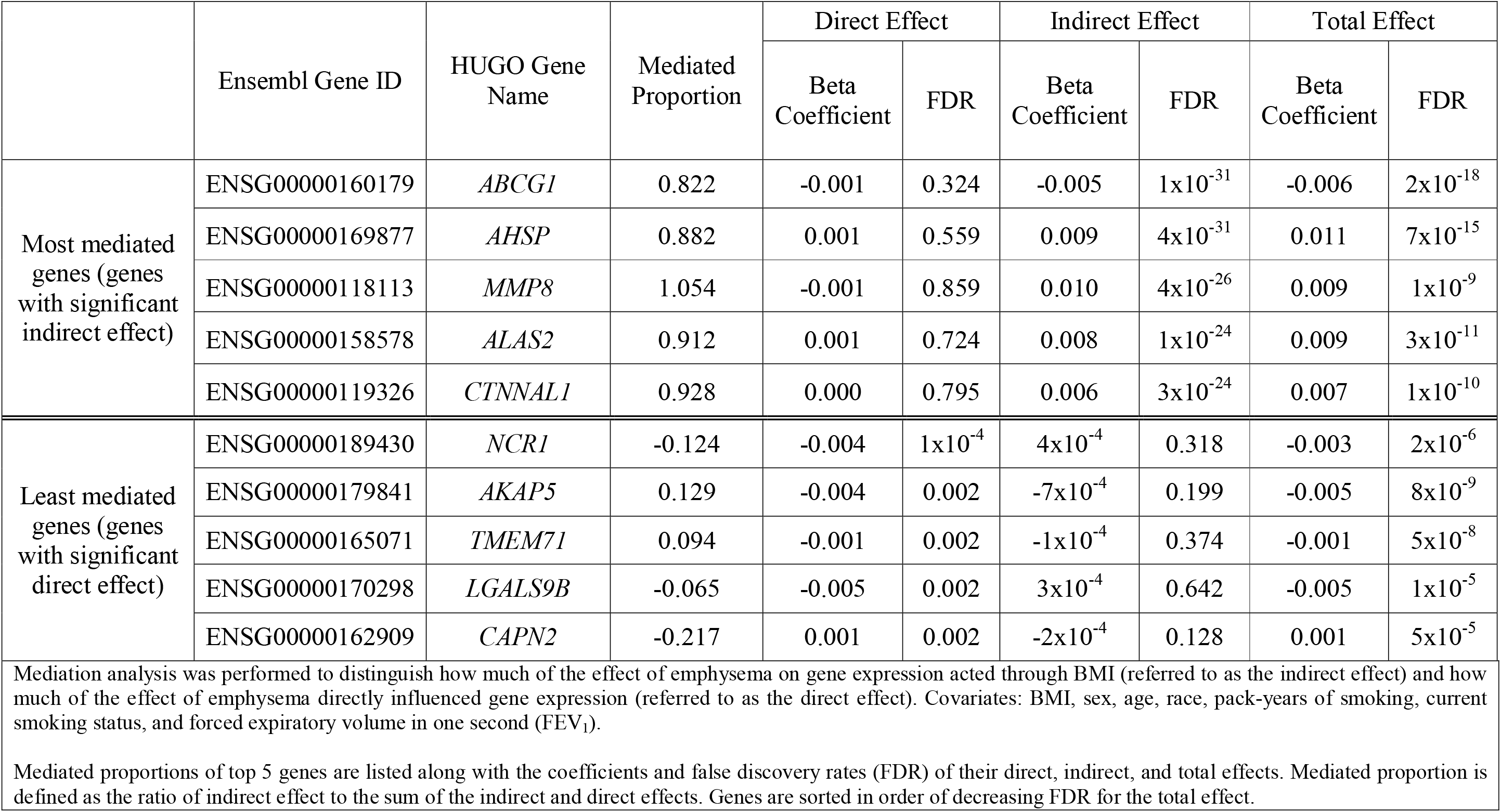
Mediated proportions and direct, indirect, and total effects of the top 5 most and least mediated differentially expressed genes significantly associated to adjusted Perc15 density.

**Figure 2.**
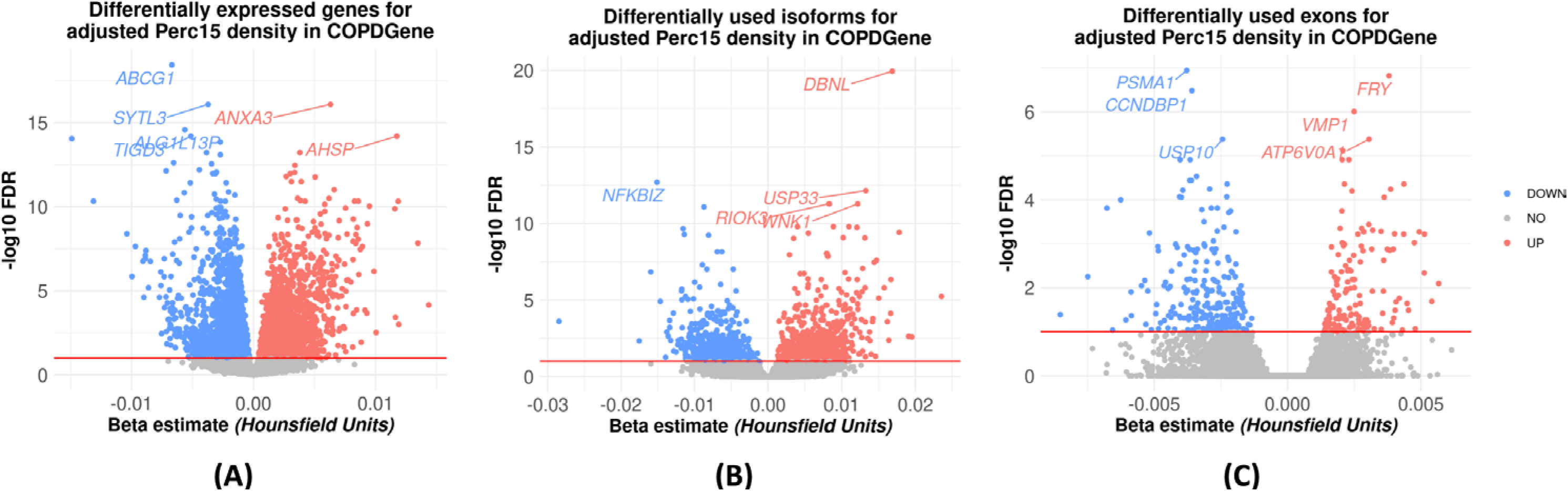
Volcano plots of the primary model representing (A) differentially expressed genes (B) differentially used isoforms, and (C) differentially used exons. Genes significantly associated with adjusted Perc15 density appear above the red line marked at FDR 10%. Up-regulated genes are in blue and down-regulated genes are in red. Isoforms/exons that are not differential used are gray and appear below the threshold line. Adjusted Perc15 density: Hounsfield units at the 15^th^ percentile of CT density histogram at total lung capacity, corrected for the inspiratory depth (per convention, adjusted Perc15 density values are reported as HU + 1000). The lower the Perc15 values are, the more CT-quantified emphysema is present. Upregulated versus downregulated genes are reported with respect to adjusted Perc15 density (i.e., they have opposite directions for their associations with emphysema).

### Differential isoform and exon usage analyses

We next performed differential isoform usage (DIU) and differential exon usage (DEU) analyses on the training dataset to investigate the changes in relative isoform and exon levels within single parent genes. Out of 78,837 isoforms and 209,707 exons tested, 1,478 isoforms and 368 exons reached significance *(FDR 10%)* (Table 2). The differentially used isoforms mapped to 1,209 individual genes (Table E4), 45% of which (542/1,209) were also identified in the DGE analysis. The differentially used exons mapped to 251 genes (Table E5), 68% of which (171/251) were also differentially expressed. 53% (788/1478) of the significant isoforms and 66% (244/368) of the significant exons were up-regulated with respect to adjusted Perc15 density (Figure 2B, 2C). The GO enrichment analyses performed on the differentially used isoforms and differentially used exons yielded 35 and 13 significantly enriched biological processes, respectively. Top processes included mitophagy, regulation of NF-κB signaling, negative regulation of WNT signaling, and viral transcription (Table 3, E6 E7).

### Protein association analysis

We tested 4,979 SomaScan proteins measured in the training dataset using multivariable linear regression. 18% (881/4,979) were significantly associated with emphysema *(FDR 10%)* (Table E8, Figure E2). Seventeen significantly enriched biological processes were identified, including pathways related to classical complement pathway and WNT signaling (Table E9). Figure 3 summarizes the overlap of the biomarkers and GO terms between DGE, DIU, DEU, and protein analyses, showing that most of the significant biomarkers and enriched pathways are unique to each analysis.

**Figure 3.**
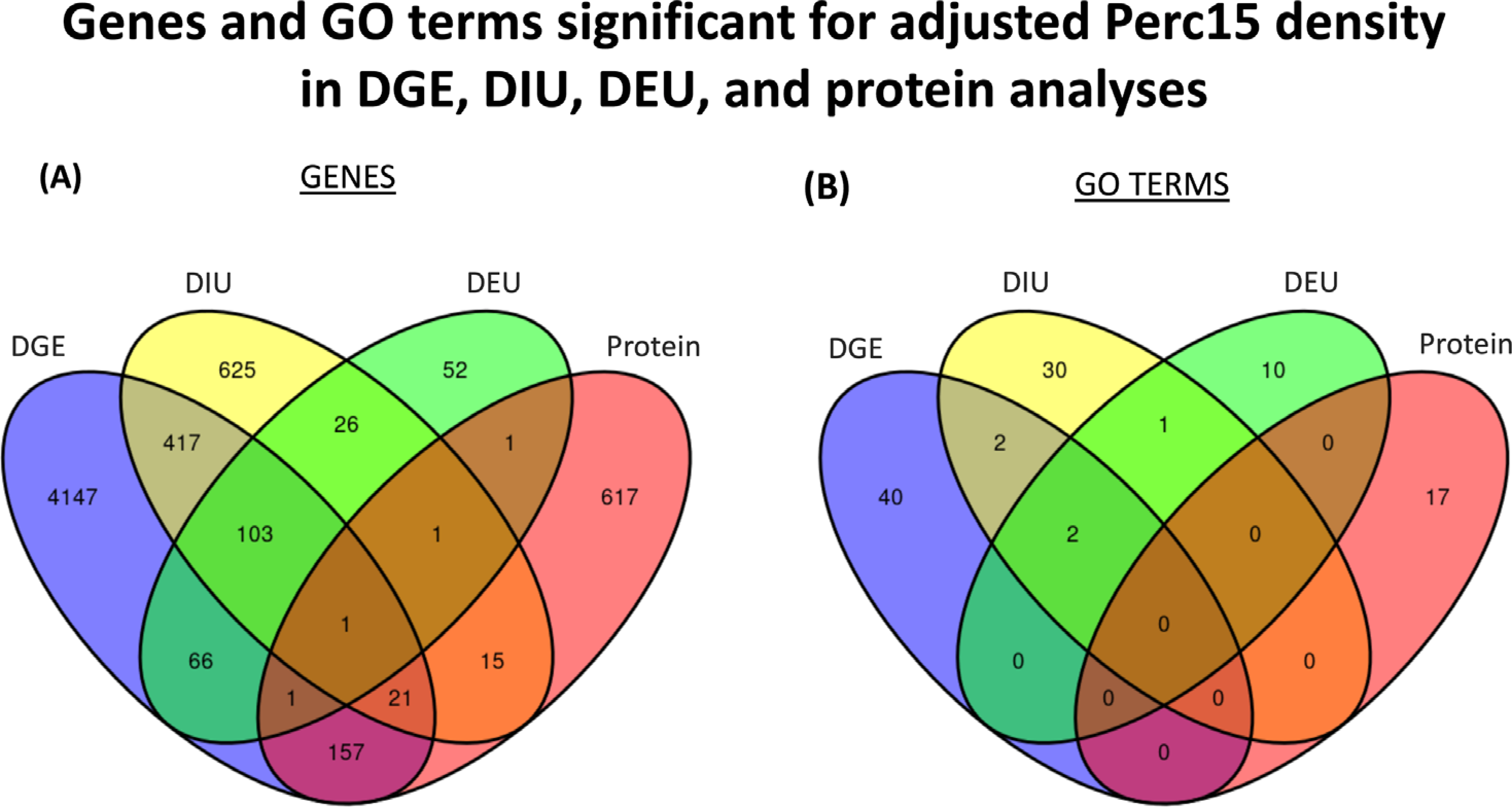
(A) Number of significant genes associated with adjusted Perc15 density from the differential gene expression (DGE), differential isoform usage (DIU), differential exon usage (DEU), and protein association analyses. HUGO gene symbols were used to find the intersection of biomarkers between the DGE, DIU, DEU, and protein analyses. Multiple proteins may map to a single gene. Therefore, the diagram does not reflect the total number of proteins significantly associated with adjusted Perc15 density. (B) Number of significant enriched gene ontology (GO) terms from the DGE, DIU, DEU, and protein association analyses. Adjusted Perc15 density: Hounsfield units at the 15^th^ percentile of CT density histogram at total lung capacity, corrected for the inspiratory depth (per convention, adjusted Perc15 density values are reported as HU + 1000). The lower the Perc15 values are, the more CT-quantified emphysema is present. Upregulated versus downregulated are reported with respect to adjusted Perc15 density (i.e., they have opposite directions for their associations with emphysema).

### Validation

We analyzed 1,016 subjects with RNA-seq and proteomic data in the testing sample to provide independent validation of the emphysema biomarkers identified in the training sample. We observed that the effect sizes were highly correlated between the analyses performed in the training and testing data for the DGE, DEU, and protein analyses (Pearson’s r = 0.80, 0.86, and 0.88, respectively). A lower correlation (r = 0.29) was observed in the DIU analysis. We further determined whether biomarkers were validated by using a threshold of (testing) P-value < 0.1 and checking if the training and testing data had a consistent direction of effect. Respectively, 46% (2,252/4,913), 30% (449/1,478), 60% (233/368), and 47% (416/881) of the DGE, DIU, DEU, and protein biomarkers were validated (Tables E2, E4, E5, and E8).

### Mediation analysis

Since severe emphysema is often associated with low BMI, we performed sensitivity analyses that also adjusted for BMI. We observed that 96% (4,728/4,913) of the genes and 80% (703/881) of the proteins (Figure E3) associated with emphysema from the primary analysis were no longer significant after adjustment for BMI (Tables E10 and E11). These observations suggest that BMI mediates many of the emphysema-associated transcriptomic and proteomic changes. To investigate this, we performed mediation analysis based on the directed acyclic graph (DAG) (Figure E4) to divide each observed biomarker association into a direct pathway (emphysema directly affects gene/protein expression) and an indirect pathway (emphysema affects gene/protein expression via its effects on BMI). Of the 4,913 differentially expressed genes and 881 proteins that reached significance in the primary model (i.e., no BMI adjustment), 70% of genes (3,456/4,913) and 61% of proteins (537/881) showed evidence of mediation with a significant indirect effect and no significant direct effect *(FDR 10%)* (Tables E12 and E13).

### Prediction

To select blood biomarkers for inclusion in cross-sectional predictive models for emphysema, we performed association analyses in the training dataset, adjusting only for technical factors, which yielded 13,066 genes, 4,254 isoforms, 2,263 exons, and 1,719 proteins that were used as candidate predictors. To evaluate whether RNA-seq is more informative at the gene, isoform, or exon level, we trained three separate models (clinical + gene, clinical + isoform, and clinical + exon). The AUROCs were 0.86, 0.74, and 0.86, respectively (Table E14 and Figure E5). Although no statistical significance was attained, a slightly higher AUROC was achieved with the genes compared to exons. Accordingly, we focused on gene-level quantifications for the subsequent models. We next evaluated the relative contribution of CBC proportions, genes, and proteins compared to a baseline model using clinical variables alone. The model using only clinical variables explained 35% of the variance of emphysema. Substantial improvement was seen from adding gene and protein data respectively (R^2^ = 0.38 for clinical + CBC + gene and 0.50 for clinical + CBC + protein, Table E14). The model with clinical + CBC + gene + protein data did not perform as well as the model with clinical + CBC + protein data. The same pattern was seen when we evaluated model performance for distinguishing subjects in the top versus bottom emphysema tertile; the highest-performing model was the clinical + CBC + protein model with an AUROC of 0.92. Figure 4 summarizes the model results, and Table E14 summarizes the AUROCs, alphas, and L1 ratios.

**Figure 4.**
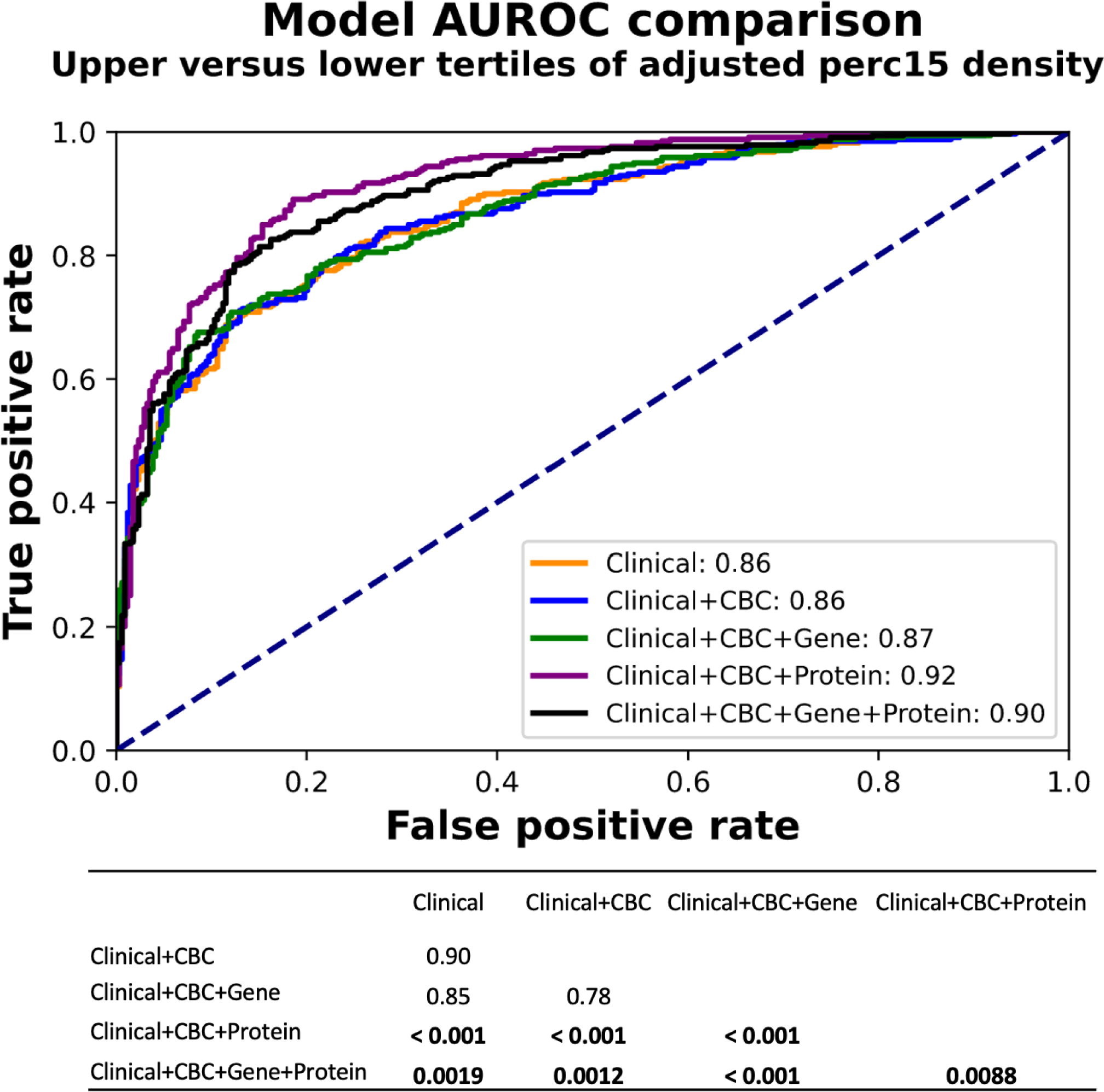
The receiver operating characteristic curves for the elastic net prediction models: clinical (age, race, sex, BMI, pack-years of smoking, and current smoking status) only, clinical + complete blood count (CBC) proportions of neutrophils, eosinophils, monocytes, lymphocytes, and platelets, clinical + CBC + genes, clinical + CBC + proteins, and clinical + CBC + genes + proteins. The table summarizes the pairwise DeLong P-values of the model comparisons. P-values < 0.05 are bolded.

Ranked by absolute beta coefficients, the top-10 predictors of the all-inclusive (clinical + CBC + gene + protein) model included BMI, sRAGE, and two biomarkers that have not been previously linked to emphysema (the *MIR124-1HG* gene and the PSMP protein) (Figure 5).

**Figure 5.**
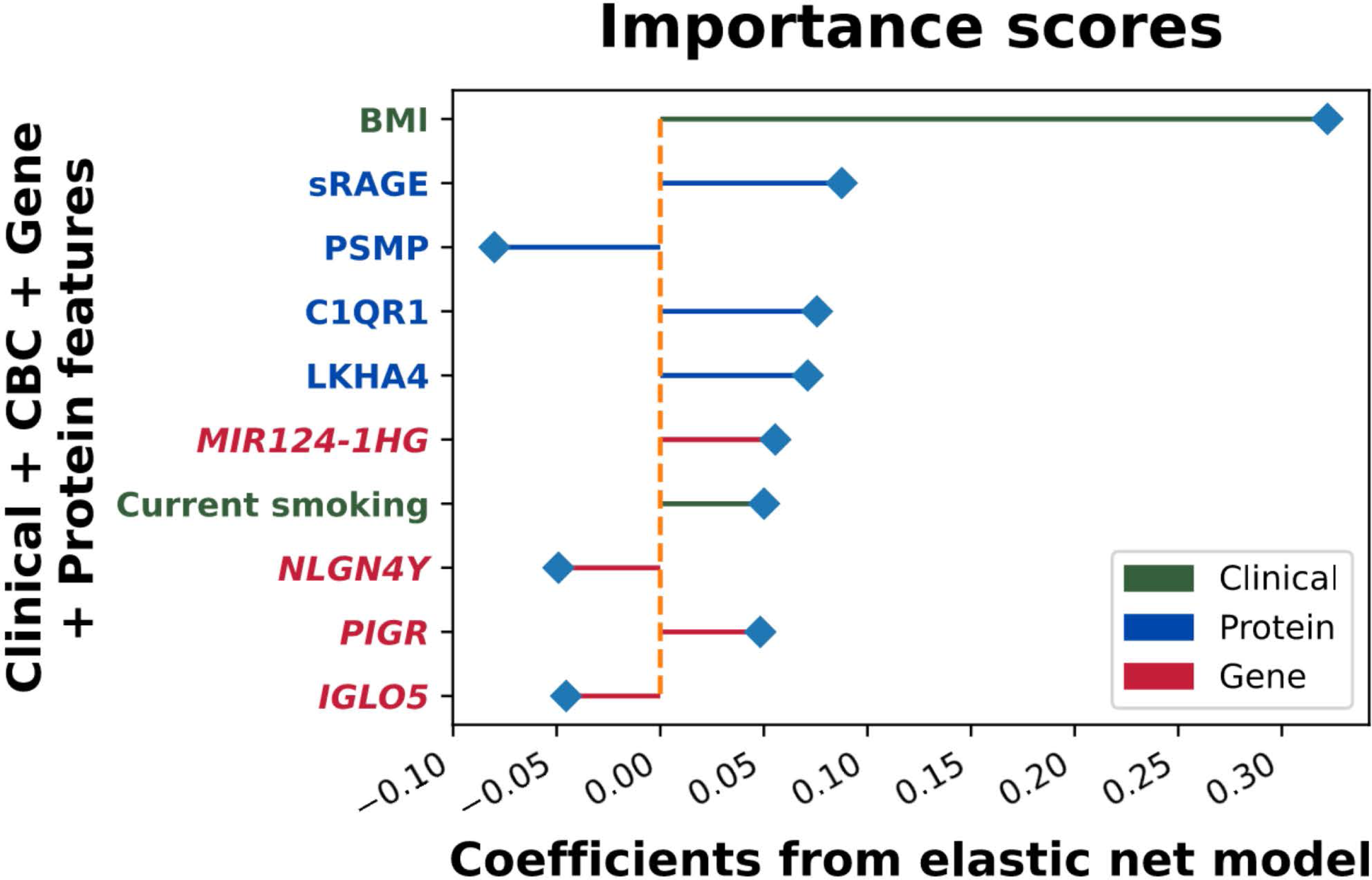
Top 10 predictors sorted in descending order by the absolute values of their beta-coefficients from the elastic net model using clinical (age, race, sex, BMI, pack-years of smoking, and current smoking status), complete blood count (CBC) proportions of neutrophils, eosinophils, monocytes, lymphocytes, and platelets, gene, and protein data. The horizontal lines represent the magnitude of the coefficient for each feature. All predictors were centered and scaled.

## DISCUSSION

In this study, we performed the largest blood transcriptomic and proteomic profiling of CT-quantified emphysema to date, including investigations into alternative splicing mechanisms, identifying thousands of validated blood biomarker associations. The biological relevance of these findings was assessed through GO pathway analyses, which mostly demonstrated enrichment for inflammatory pathways and cell differentiation. Mediation analysis revealed that 70% of the differentially expressed genes and 61% of the associated proteins are mediated through BMI, implying that blood biomarker associations to emphysema largely reflect shared biological processes with BMI. We also demonstrated the utility of incorporating multi-omics data in enhancing the prediction of emphysema.

In previous biomarker studies of emphysema, the extracellular matrix (ECM), NF-κB, transforming growth factor beta (TGF-ß), B cell antigen receptor (BCR), and oxidative phosphorylation pathways were among the most frequently reported emphysema-associated pathways (8, 46, 47). However, most of these studies focused on a single ‘omics modality (20, 24, 48). Our investigation of blood-based transcriptomic and proteomic biomarkers supported numerous established emphysema-associated pathways and discovered new ones. In addition, our alternative splicing analysis for the first time revealed widespread evidence of alternative splicing associated with emphysema.

Most blood biomarker associations with emphysema occur through BMI, as indicated by the significant mediation of the tested genes and proteins. This suggests that some of the molecular processes identified in this analysis may be causally related to both emphysema and BMI. We must keep in mind, though, that our mediation analysis is based on the following assumptions: no unmeasured confounding of the emphysema-BMI-gene expression/protein level relationship, no measurement error for the exposure or mediator, and the arrows in the DAG are correctly specified. While our specified DAG is reasonable based on prior knowledge, there are other plausible alternatives DAGs, but no currently available methods to simultaneously test these possibilities.

CT scan is the best non-invasive method for detecting emphysema. However, CT has several drawbacks, including increased costs, radiation exposure, and high rates of unrelated false-positive findings (5). Accurate risk prediction tools that use the best available data sources to stratify patients based on their specific risk profiles could help with more efficient early and targeted interventions. Until recently, such prediction models were only created using data from a single ‘omics type with or without standard clinical features (49-52). As the first study to utilize genes, alternative splicing, and proteins combined with clinical and CBC predictors, we developed models that could classify upper and lower tertiles of emphysema severity with good accuracy. While alternative splicing predictors were worth exploring, gene data had a higher AUROC. While genes outperformed clinical and CBC features, protein predictors yielded the best AUROC across all models.

From the top 10 predictors of the clinical + CBC + gene + protein model, sRAGE, which minimizes tissue injury and inflammation, has consistently been recognized as a candidate emphysema biomarker (5, 10, 53, 54). Not previously connected to emphysema, PSMP has been implicated in inflammation and cancer development (55, 56) and *MIR124-1HG* has been shown to affect Wnt-signaling and inflammation (57-60).Their putative roles and functions in emphysema require further investigation.

This study has several strengths. The large sample size allowed us to identify many more significant associations than any previous study, and we could split our sample to allow for the validation of our findings. This is the first study that has examined alternative splicing mechanisms in emphysema in addition to differential gene expression and protein association analyses. Because emphysema often co-occurs with low BMI, we performed mediation analyses to better understand the relationship between molecular markers, emphysema, and BMI, providing suggestive evidence of shared biology between emphysema and BMI. Finally, we were able to improve emphysema prediction models with the use of multi-omic data.

This study also has several limitations. Complete blood count quantifications do not capture the variability of the immune cell subpopulations, which limits the ability to localize these effects to specific cell types. Future studies may address this by using single-cell transcriptomics data. Limitations to the SomaScan proteomics include the lack of SOMAmers for small molecules such as desmosine, fibrinogen degradation product (Aa-Val360, a specific product generated by elastase cleavage of fibrinogen), and sphingomyelin, which have been suggested to be emphysema biomarkers in other studies (37, 60-62). Lastly, the mediation analysis needs to be viewed as hypothesis-generating since it is based on a number of assumptions.

## CONCLUSION

Our transcriptomic and proteomic analyses yielded numerous inflammatory and cell differentiation pathways connected to emphysema as well as novel potential blood biomarkers of the disease. While not yet ready to be used in clinical practice, with further validation, our prediction model might be helpful as a less invasive indicator of emphysema severity that could guide patient enrollment in clinical trials. Future research is necessary to compare blood and lung tissue biomarkers, understand how they change as emphysema progresses, and evaluate the impact of implementing these predictive models to personalize and improve patient care.

## Data Availability

All data produced in the present study are available upon reasonable request to the authors

## COPDGene Investigators - Core Units

### Administrative Center

James D. Crapo, MD (PI); Edwin K. Silverman, MD, PhD (PI); Barry J. Make, MD; Elizabeth A. Regan, MD, PhD

### Genetic Analysis Center

Terri H. Beaty, PhD; Peter J. Castaldi, MD, MSc; Michael H. Cho, MD, MPH; Dawn L. DeMeo, MD, MPH; Adel Boueiz, MD, MMSc; Marilyn G. Foreman, MD, MS; Auyon Ghosh, MD; Lystra P. Hayden, MD, MMSc; Craig P. Hersh, MD, MPH; Jacqueline Hetmanski, MS; Brian D. Hobbs, MD, MMSc; John E. Hokanson, MPH, PhD; Wonji Kim, PhD; Nan Laird, PhD; Christoph Lange, PhD; Sharon M. Lutz, PhD; Merry-Lynn McDonald, PhD; Dmitry Prokopenko, PhD; Matthew Moll, MD, MPH; Jarrett Morrow, PhD; Dandi Qiao, PhD; Elizabeth A. Regan, MD, PhD; Aabida Saferali, PhD; Phuwanat Sakornsakolpat, MD; Edwin K. Silverman, MD, PhD; Emily S. Wan, MD; Jeong Yun, MD, MPH

### Imaging Center

Juan Pablo Centeno; Jean-Paul Charbonnier, PhD; Harvey O. Coxson, PhD; Craig J. Galban, PhD; MeiLan K. Han, MD, MS; Eric A. Hoffman, Stephen Humphries, PhD; Francine L. Jacobson, MD, MPH; Philip F. Judy, PhD; Ella A. Kazerooni, MD; Alex Kluiber; David A. Lynch, MB; Pietro Nardelli, PhD; John D. Newell, Jr., MD; Aleena Notary; Andrea Oh, MD; Elizabeth A. Regan, MD, PhD; James C. Ross, PhD; Raul San Jose Estepar, PhD; Joyce Schroeder, MD; Jered Sieren; Berend C. Stoel, PhD; Juerg Tschirren, PhD; Edwin Van Beek, MD, PhD; Bram van Ginneken, PhD; Eva van Rikxoort, PhD; Gonzalo Vegas SanchezFerrero, PhD; Lucas Veitel; George R. Washko, MD; Carla G. Wilson, MS

### PFT QA Center

Salt Lake City, UT: Robert Jensen, PhD

### Data Coordinating Center and Biostatistics

National Jewish Health, Denver, CO: Douglas Everett, PhD; Jim Crooks, PhD; Katherine Pratte, PhD; Matt Strand, PhD; Carla G. Wilson, MS

### Epidemiology Core

University of Colorado Anschutz Medical Campus, Aurora, CO: John E. Hokanson, MPH, PhD; Erin Austin, PhD; Gregory Kinney, MPH, PhD; Sharon M. Lutz, PhD; Kendra A. Young, PhD

### Mortality Adjudication Core

Surya P. Bhatt, MD; Jessica Bon, MD; Alejandro A. Diaz, MD, MPH; MeiLan K. Han, MD, MS; Barry Make, MD; Susan Murray, ScD; Elizabeth Regan, MD; Xavier Soler, MD; Carla G. Wilson, MS

### Biomarker Core

Russell P. Bowler, MD, PhD; Katerina Kechris, PhD; Farnoush BanaeiKashani, PhD

## COPDGene Investigators - Clinical Centers

### Ann Arbor VA

Jeffrey L. Curtis, MD; Perry G. Pernicano, MD

### Baylor College of Medicine, Houston, TX

Nicola Hanania, MD, MS; Mustafa Atik, MD; Aladin Boriek, PhD; Kalpatha Guntupalli, MD; Elizabeth Guy, MD; Amit Parulekar, MD

### Brigham and Women’s Hospital, Boston, MA

Dawn L. DeMeo, MD, MPH; Craig Hersh, MD, MPH; Francine L. Jacobson, MD, MPH; George Washko, MD

### Columbia University, New York, NY

R. Graham Barr, MD, DrPH; John Austin, MD; Belinda D’Souza, MD; Byron Thomashow, MD

### Duke University Medical Center, Durham, NC

Neil MacIntyre, Jr., MD; H. Page McAdams, MD; Lacey Washington, MD

### HealthPartners Research Institute, Minneapolis, MN

Charlene McEvoy, MD, MPH; Joseph Tashjian, MD

### Johns Hopkins University, Baltimore, MD

Robert Wise, MD; Robert Brown, MD; Nadia N. Hansel, MD, MPH; Karen Horton, MD; Allison Lambert, MD, MHS; Nirupama Putcha, MD, MHS

### Lundquist Institute for Biomedical Innovation at Harbor UCLA Medical Center, Torrance, CA

Richard Casaburi, PhD, MD; Alessandra Adami, PhD; Matthew Budoff, MD; Hans Fischer, MD; Janos Porszasz, MD, PhD; Harry Rossiter, PhD; William Stringer, MD

### Michael E. DeBakey VAMC, Houston, TX

Amir Sharafkhaneh, MD, PhD; Charlie Lan, DO

### Minneapolis VA

Christine Wendt, MD; Brian Bell, MD; Ken M. Kunisaki, MD, MS

### Morehouse School of Medicine, Atlanta, GA

Eric L. Flenaugh, MD; Hirut Gebrekristos, PhD; Mario Ponce, MD; Silanath Terpenning, MD; Gloria Westney, MD, MS

### National Jewish Health, Denver, CO

Russell Bowler, MD, PhD; David A. Lynch, MB

### Reliant Medical Group, Worcester, MA

Richard Rosiello, MD; David Pace, MD

### Temple University, Philadelphia, PA

Gerard Criner, MD; David Ciccolella, MD; Francis Cordova, MD; Chandra Dass, MD; Gilbert D’Alonzo, DO; Parag Desai, MD; Michael Jacobs, PharmD; Steven Kelsen, MD, PhD; Victor Kim, MD; A. James Mamary, MD; Nathaniel Marchetti, DO; Aditi Satti, MD; Kartik Shenoy, MD; Robert M. Steiner, MD; Alex Swift, MD; Irene Swift, MD; Maria Elena Vega-Sanchez, MD

### University of Alabama, Birmingham, AL

Mark Dransfield, MD; William Bailey, MD; Surya P. Bhatt, MD; Anand Iyer, MD; Hrudaya Nath, MD; J. Michael Wells, MD

University of California, San Diego, CA: Douglas Conrad, MD; Xavier Soler, MD, PhD; Andrew Yen, MD

### University of Iowa, Iowa City, IA

Alejandro P. Comellas, MD; Karin F. Hoth, PhD; John Newell, Jr., MD; Brad Thompson, MD

### University of Michigan, Ann Arbor, MI

MeiLan K. Han, MD MS; Ella Kazerooni, MD MS; Wassim Labaki, MD MS; Craig Galban, PhD; Dharshan Vummidi, MD

### University of Minnesota, Minneapolis, MN

Joanne Billings, MD; Abbie Begnaud, MD; Tadashi Allen, MD

### University of Pittsburgh, Pittsburgh, PA

Frank Sciurba, MD; Jessica Bon, MD; Divay Chandra, MD, MSc; Joel Weissfeld, MD, MPH

### University of Texas Health, San Antonio, San Antonio, TX

Antonio Anzueto, MD; Sandra Adams, MD; Diego Maselli-Caceres, MD; Mario E. Ruiz, MD; Harjinder Singh, MD

