## Supplement for "Blood-Based Transcriptomic and Proteomic Biomarkers of Emphysema"

Rahul Suryadevara^1^, Andrew Gregory^1^, Robin Lu^1^, Zhonghui Xu^1^, Aria Masoomi^2^, Sharon M. Lutz^3^, Seth Berman^1^, Jeong H. Yun^1,4^, Aabida Saferali^1^, Craig P. Hersh^1,4^, Edwin K. Silverman^1,4^, Jennifer Dy^2^, Katherine A. Pratte^5^, Russell P. Bowler^5^, Peter J. Castaldi^1,6^*, Adel Boueiz^1,4^* for the COPDGene investigators.

* Equal contribution

^1^Channing Division of Network Medicine, Brigham and Women’s Hospital, Harvard Medical School, Boston, MA; ^2^Department of Electrical and Computer Engineering, Northeastern University, Boston, MA; ^3^Department of Population Medicine, Harvard Pilgrim Health Care Institute, Boston, MA; ^4^Division of Pulmonary and Critical Care Medicine, Brigham and Women’s Hospital, Harvard Medical School, Boston, MA; ^5^Department of Biostatistics, National Jewish Health, Denver, CO; ^5^Division of Pulmonary, Critical Care and Sleep Medicine, National Jewish Health, Denver, CO; ^6^Division of General Medicine and Primary Care, Brigham and Women’s Hospital, Harvard Medical School, Boston, MA.

**Supplemental methods**

***RNA extraction, sequencing, and expression quantification***

Total blood RNA was extracted from subjects at Visit 2 in PAXgene TM Blood RNA tubes from the Qiagen PreAnalytiX PAXgene Blood miRNA Kit (Qiagen, Valencia, CA). Paired-end reads were generated from Illumina sequencers and aligned to the GRCh38 genome using STAR (1). Gene transfer format (GTF) annotation was downloaded from the Biomart Ensembl database (Ensembl Genes release 94, GRCh38.p12 assembly) on October 21, 2018. Exons from the GTF were broken into disjoint parts (exonic parts) (2). Sequencing read counts on exonic parts were generated from featureCounts in Rsubread (v1.32.2) (3). Isoform and gene expression counts were obtained using Salmon (v0.12.0) and tximport (v1.10.0) (4, 5). The gene and isoform count data used for this analysis are available in the Gene Expression Omnibus (6, 7) (accession number GSE158699).

***Mediation analysis***

Using the covariates from the emphysema model without BMI adjustment, we fitted a natural effect model, which models both direct and indirect effects, using the imputation approach implemented in the medflex package (8). Due to collinearity that may bias the mediation analysis, we excluded the library preparation batch, CBC, and scanner model from the mediation analysis model, leaving only the BMI, sex, age, race, pack-years of smoking, current smoking status, and FEV_1_ as covariates.

***Elastic net***

Elastic net offers several well-known benefits, including the ability to account for multi-collinear features and avoid overfitting (9). Elastic net is a regression model that combines the L1 and L2 penalty functions. The L1 penalty shrinks features’ coefficients to zero, while the L2 penalty adds a penalty equivalent to the square of the magnitude of the features’ coefficients (10, 11).

$$\hat{\beta}\equiv\mathrm{argmin}_{\beta}(\left\| y-X\beta\right\|^{2}+\lambda_{2}\left\| \beta\right\|^{2}+\lambda_{1}\left\| \beta\right\|_{1})$$

Given data input X and its labels y, elastic net fits a linear model between X and y by learning the 𝛽 coefficient such that the model is a good linear predictor of y, but also the norm L1 (encourages sparsity) and L2 (preventing coefficients from becoming too large) are penalized (10).

We used the Python implementation of the elastic net from the scikit-learn (v1.0.1), which takes in alpha and L1 ratio parameters. For reproducibility, we initialized all elastic net models at random state = 0. The models were tuned using 10-fold cross-validation, and GridSearchCV from the scikit-learn Python package was utilized for hyperparameter tuning. Specifically, the alpha and L1 ratio parameters were tuned for each model to accommodate for varying feature sizes. The best score from the grid search was determined using negative mean absolute error. The elastic net models were fitted and trained using the ideal alpha and L1 ratio parameters identified by the grid search.

**Supplemental Figure Legends**

**Figure E1.** Study overview figure. Abbreviations: BMI = Body mass index, CBC = Cell blood count, CT = Computed tomography.

**Figure E2.** Forest plot of beta estimates for the top 15 plasma proteins associated with adjusted Perc15 density, ranked by false discovery rate. The point represents the beta estimate for adjusted Perc15, measured in Hounsfield Units. A 95% confidence interval surrounds each point. Adjusted Perc15 density: Hounsfield units at the 15^th^ percentile of CT density histogram at total lung capacity, corrected for the inspiratory depth (per convention, adjusted Perc15 density values are reported as HU + 1000). The lower the Perc15 values are, the more CT-quantified emphysema is present. Positive *versus* negative directions of the protein coefficients are reported with respect to the associations with adjusted Perc15 density (i.e., they have opposite directions for their associations with emphysema).

**Figure E3.** Log fold-change *versus* average log-expression of genes from the differential gene expression (DGE) analysis for adjusted Perc15 density without BMI adjustment (A) and with BMI adjustment (B). Blue represents differentially expressed genes at FDR 10%*.* Black represents genes that did not reach statistical significance. Volcano plot of proteins without BMI adjustment (C) and with BMI adjustment (D). Proteins significantly associated with adjusted Perc15 density appear over the red line marked at FDR 10%. Proteins that are significantly associated with emphysema are marked blue and those that are not marked black. Adjusted Perc15 density: Hounsfield units at the 15^th^ percentile of CT density histogram at total lung capacity, corrected for the inspiratory depth (per convention, adjusted Perc15 density values are reported as HU + 1000). The lower the Perc15 values are, the more CT-quantified emphysema is present. Upregulated *versus* downregulated genes are provided with respect to their relationships with adjusted Perc15 density (i.e., they have opposite directions for their associations with emphysema).

**Figure E4.** Directed acyclic graph (DAG) representing the direction of effects between emphysema, gene expression or protein levels, and BMI. The direct effect is represented by the arrow pointing from emphysema to gene expression or protein levels. The indirect effect goes in the same direction, through BMI. The total effect is the sum of the direct and indirect effects.

**Figure E5.** The receiver operating characteristic curves for the following models from the elastic net prediction: Clinical (age, race, sex, BMI, pack-years of smoking, and current smoking status) only, clinical + gene, clinical + isoform, and clinical + exon. The table insert summarizes the pairwise DeLong P-values of the models, in which significant differences in model performance are bolded.
