## Supplementary figures for "Blood-Based Transcriptomic and Proteomic Biomarkers of Emphysema"

### Figure E1

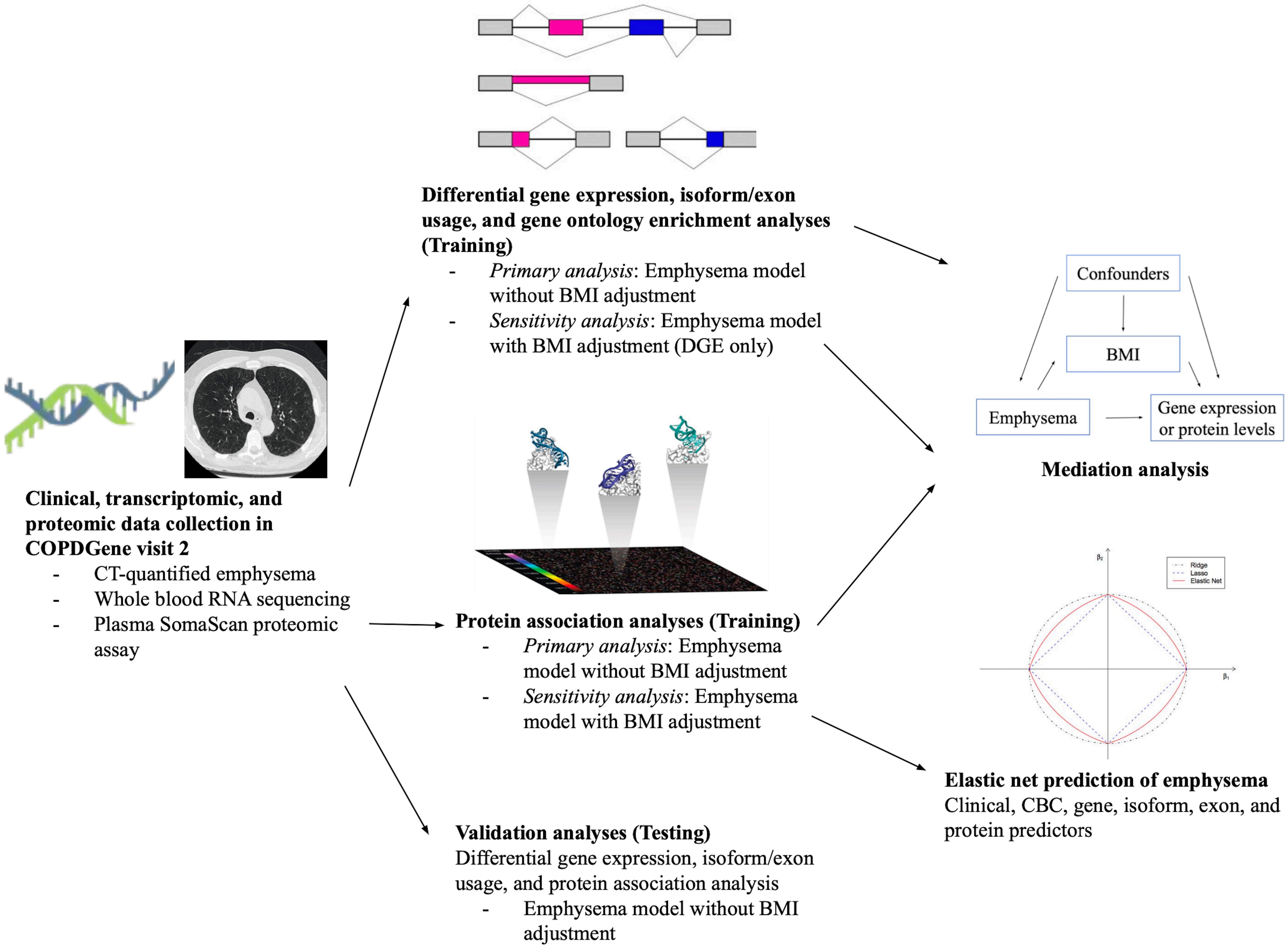
Differential gene expression, isoform/exon usage, and gene ontology enrichment analyses (Training)
- Primary analysis: Emphysema model without BMI adjustment
- Sensitivity analysis: Emphysema model with BMI adjustment (DGE only)
Confounders
BMI
Emphysema
Gene expression or protein levels
Mediation analysis
Clinical, transcriptomic, and proteomic data collection in COPDGene visit 2
- CT-quantified emphysema
- Whole blood RNA sequencing
- Plasma SomaScan proteomic assay
Protein association analyses (Training)
- Primary analysis: Emphysema model without BMI adjustment
- Sensitivity analysis: Emphysema model with BMI adjustment
Ridge
Lasso
Elastic Net
Elastic net prediction of emphysema
Clinical, CBC, gene, isoform, exon, and protein predictors
Validation analyses (Testing)
Differential gene expression, isoform/exon usage, and protein association analysis
- Emphysema model without BMI adjustment

### Figure E2

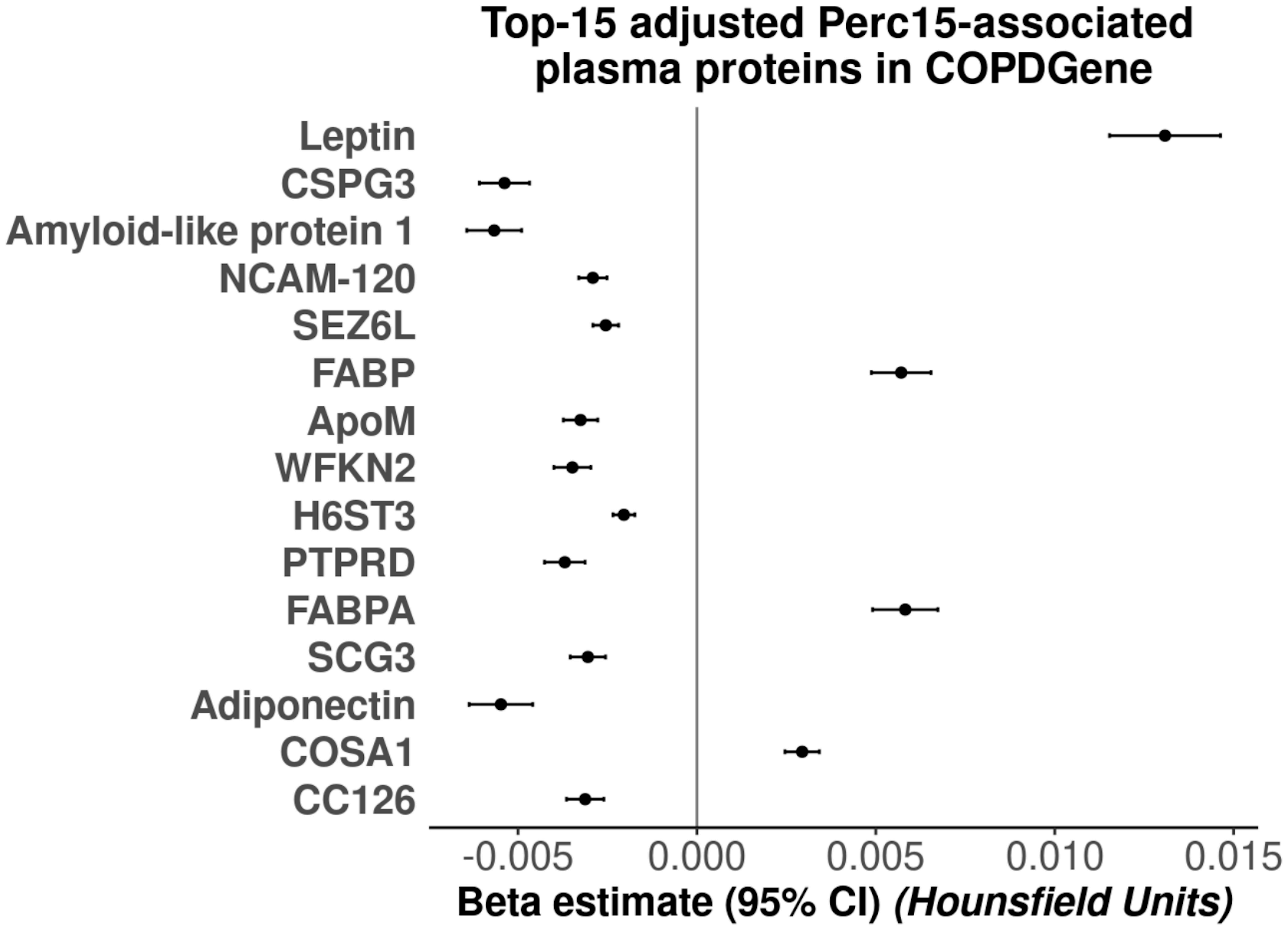
Top-15 adjusted Perc15-associated plasma proteins in COPDGene
Leptin
CSPG3
Amyloid-like protein 1
NCAM-120
SEZ6L
FABP
ApoM
WFKN2
H6ST3
PTPRD
FABPA
SCG3
Adiponectin
COSA1
CC126
-0.005
0.000
0.005
0.010
0.015
Beta estimate (95% CI) (Hounsfield Units)

### Figure E3

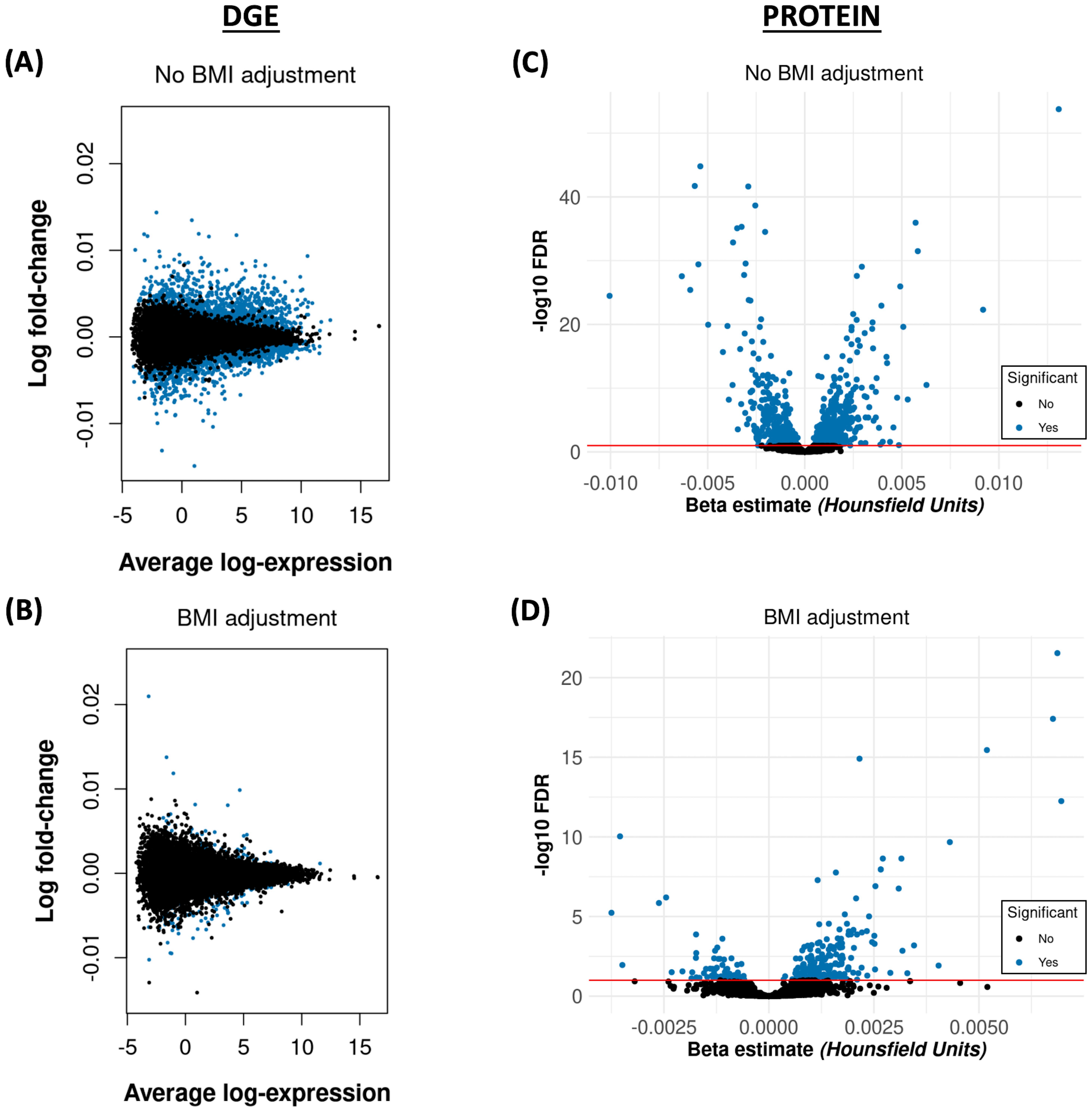
DGE
PROTEIN
(A)
No BMI adjustment
Log fold-change
Average log-expression
(B)
BMI adjustment
Log fold-change
Average log-expression
(C)
No BMI adjustment
-log10 FDR
Beta estimate (Hounsfield Units)
Significant
No
Yes
(D)
BMI adjustment
-log10 FDR
Beta estimate (Hounsfield Units)
Significant
No
Yes

### Figure E4

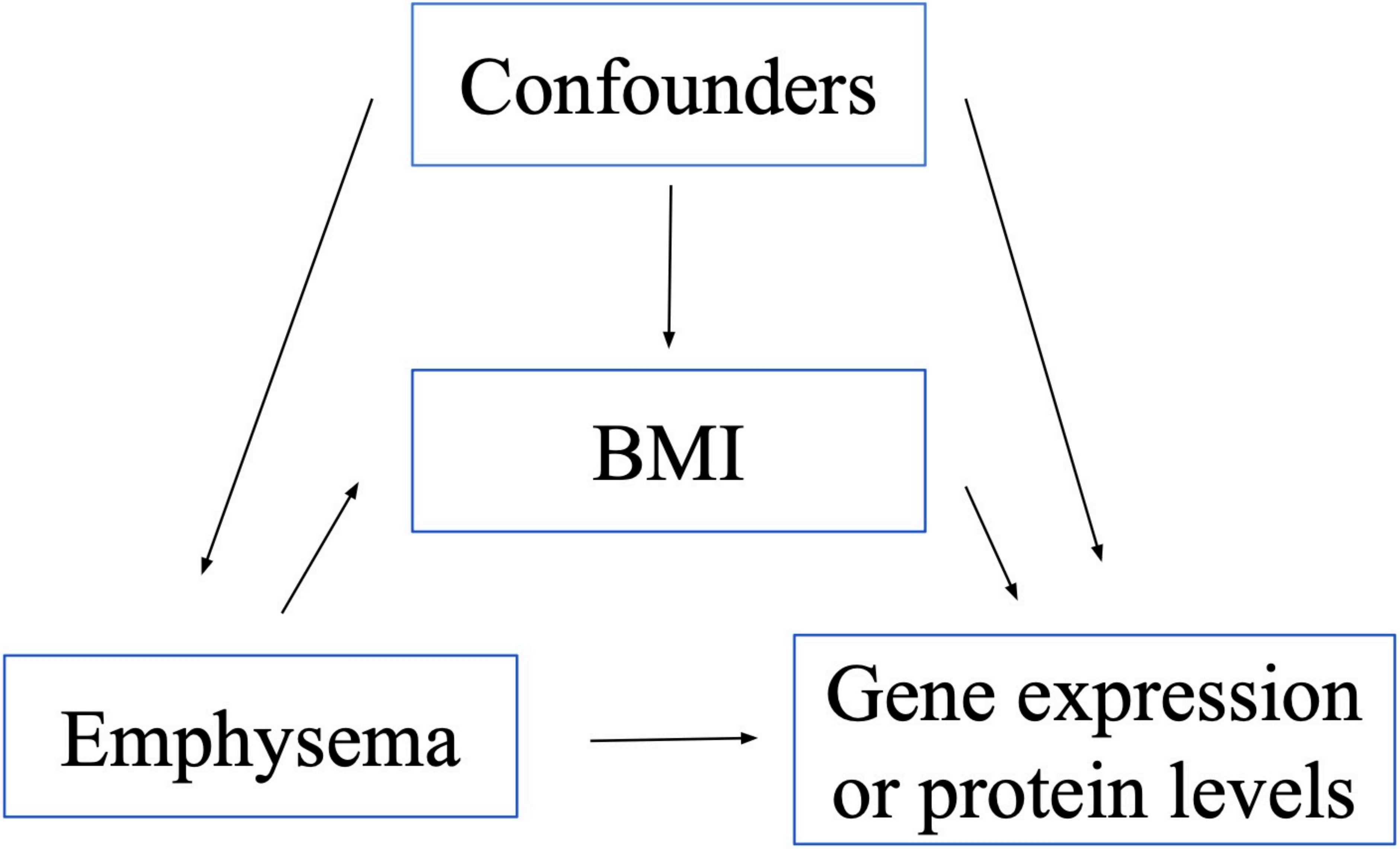
Confounders
BMI
Emphysema
Gene expression
or protein levels

### Figure E5

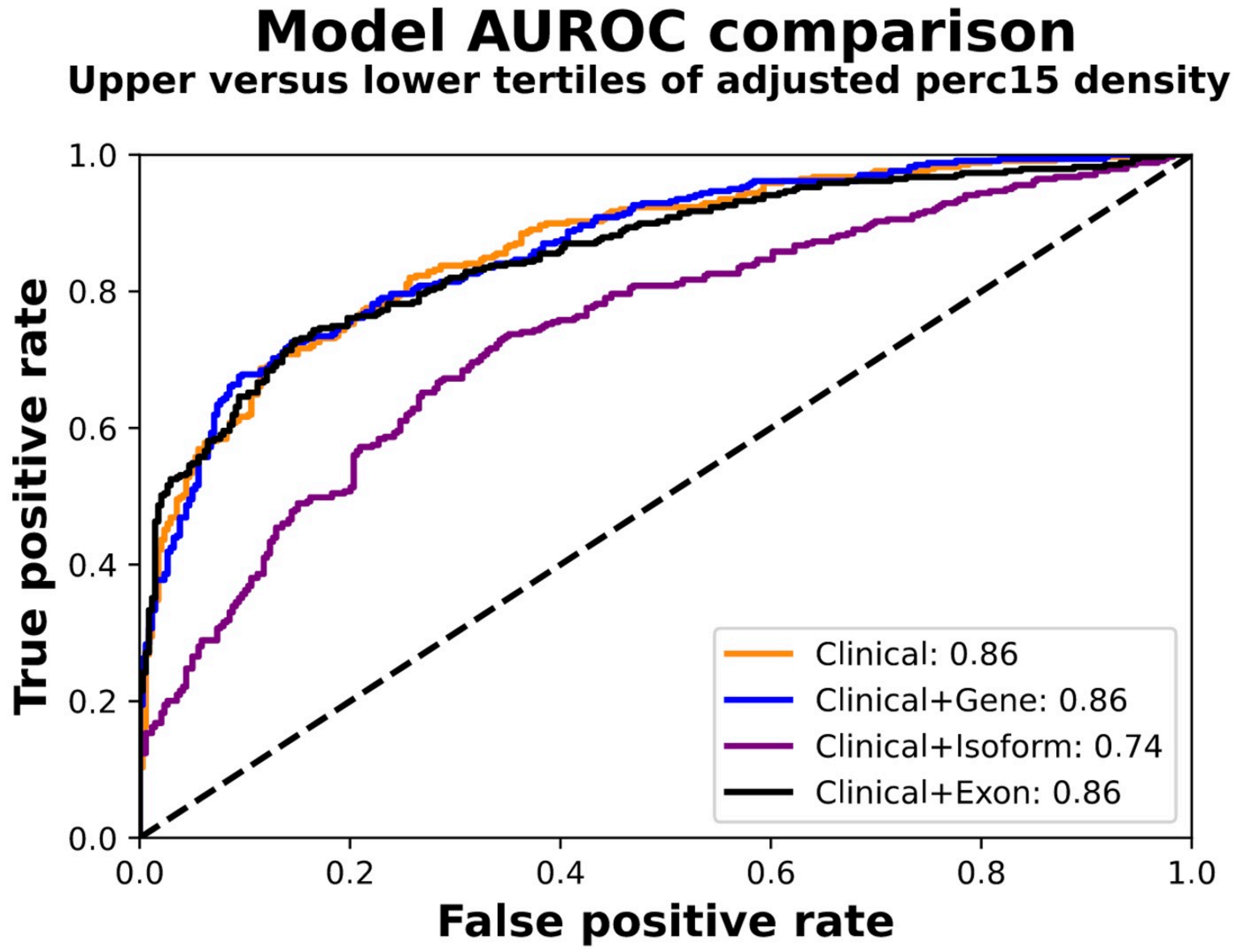


| | Clinical | Clinical+Gene | Clinical+Isoform |
|---|---|---|---|
| Clinical+Gene | 0.97 | | |
| Clinical+Isoform | **< 0.001** | **< 0.001** | |
| Clinical+Exon | 0.26 | 0.27 | **< 0.001** |
